# Infection spread simulation technology in a mixed state of multi variant viruses

**DOI:** 10.1101/2021.06.28.21259679

**Authors:** Makoto Koizumi, Motoaki Utamura, Seiichi Kirikami

## Abstract

ATLM was extended to simulate the spread of infection in a mixed state of mutant virus and conventional virus. It is applied to the 4th wave of infection spread in Tokyo, and (1) the 4th wave bottoms out near the end of the state of emergency, and the number of infected people increases again. (2) The rate of increase will be mainly by L452R virus, while the increase by N501Y virus will be suppressed. (3) It is anticipated that the infection will spread during the Olympic Games. (4) When mutant virus competes, the infection of highly infectious virus rises sharply while the infection by weakly infectious ones has converged. (5) It is effective as an infection control measure to find an infected person early and shorten the period from infection to quarantine by PCR test or antigen test as a measure other than vaccine.

## 1. INTRDUCTION

Mutation of SARS-CoV-2 that have occurred in the spread areas of the United Kingdom, Brazil, and India over the past year have also entered Japan. Mutant viruses include those with weakened infectivity and those with increased infectivity. The problem is epidemic under a virus with increased infectivity and a conventional virus coexist. In this case, modified viruses are replaced the conventional type and larger spread is expected. Therefore, the means for suppress of infection spread is important. SIR [1] and SEIR [2] models are known as simulation models for the spread of infection. Because of the simplicity from mathematical structure of models and the short calculation time, these models are often used in early infection stage. Kuniya et al. use SEIR to evaluate the effect of the state of emergency in the second wave in Tokyo [3]. Now that the infection has progressed considerably, there is a need for comprehensive simulation technology that incorporates the effects of vaccines and lockdowns. For this as well, a simulation that incorporates these effects into the coefficients is also performed at SEIR [3]. B Ritton et al. applied improved SEIR to spread infection under the case of non-uniform population structure [4]. On the other hand, ABM (Agent Based Model) has been developed [5] [6] [7] [8] [9]. This technology is a probabilistic method, and unlike the deterministic methods such as SIR and SEIR, it is a method that assumes various behaviors of a person and calculates the infection probability, and it takes a considerable amount of calculation time. The above methods do not have a time delay from infection to quarantine. We considered that the time required until isolated from infected is important role of contribution in expanding infection, therefore we developed ATLM (Apparent Time Lag Model) with delay until isolation time[10]. This model currently has an extended version with vaccine and lockdown effects [11]. The infectivity of mutant viruses has already been reported [12] and the spread of the virus has been simulated. However, there are few knowledges about infection patterns by multi variant viruses and it has not been understood yet how each mutation virus to spread or to contribute, also how strong mutation virus to become mainstream spread. In this report, we have expanded it to handle mutant viruses.

## 2. ANALISIS MODEL

If there is only one type of infectivity, several proposals such as SIR and SEIR have been proposed. The ATLM we have developed uses the following equation, which takes into account the time delay from infection to quarantine and the time delay from infection to loss of infectivity.

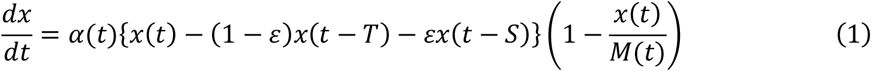

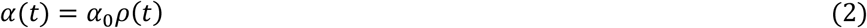

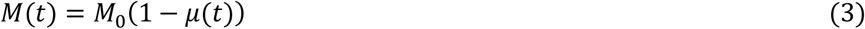

where, *x*: cumulative number of infected persons, *T*: number of days from infection to quarantine, *μ(t)*: vaccination rate, α: infection coefficient, *ε*: ratio of asymptomatic persons, *S* is the number of days from infection to extinction of infectivity. M indicates the sensitive population. ρ (t) is the rate of decrease in the infection coefficient due to the restriction of human flow due to lockdown. Equation (3) represents the decrease in the target population due to vaccination. To extend the above equation for handling the modified viruses, the following assumptions are taken into account.

1. Infected persons are infected with only one type of virus, and there is no simultaneous infection.
2. Patients who have been infected with one mutant virus in the past are not infected with other mutant virus.
3. The infection rates between viruses are independent of each other and do not interfere with each other.
4. The effect of the vaccine is the same for each mutant will.
5. The time until the onset and the time until the infectivity disappears are the same.

Above assumptions the group, *i* species modified viruses formula spread by is as follows.

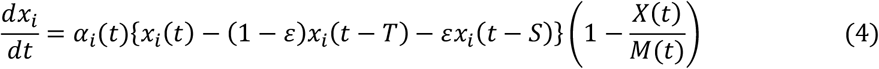

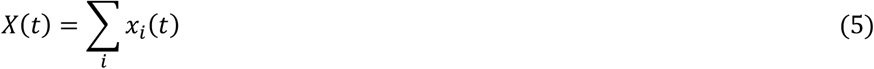

Equation (5) is a limitation that comes from assumptions (1) and (2). That is, it is shown that the infected target is the same susceptible population even if the mutant species are different.

## 3. TIME INTEGRAION

The analytical solution of the differential equation of Eq. (4) is unknown. Therefore, to solve the above equation, the 4th-order Runge-Kutta method was used for numerical integration. In equation (4). *t-T* and *t-S* value point of *x*_*i*_ in the calculation time t, is computed, especially no problem on accuracy. However, the calculation start time of the point is tricky because it is unknown. Therefore, the initial value is given a sufficiently small value compared to M_0_. Next, it should be noted that, when *X(t)* is increased, too close to *M(t)*. Especially when the vaccination rate becomes high, *X(t)* >*M(t)* may occur, in which case the solution oscillates and becomes unstable. Here, if *X(t)*/*M(t)*>1, we set the righthand side of equation (4) equal zero.

## 4. APPLICATION TO THE 4TH WAVE OF INFECTION SPREAD IN TOKYO

### 4.1 Analysis conditions

Mutant virus that is prevalent in Tokyo and is seen as almost by N501Y virus [12]. Currently, the virus of interest is L452R virus found in India. It is confirmed that 9 people are infected at 2021/ 5 / 31 by L452R virus. Investigation of mutant virus of infected persons 10 is sampling percent, therefore about 90 people are infected [13]. For infectivity, N501Y virus prevalent at this stage of infectivity is believed as 1.32 times that of the conventional virus type [12]. L452R virus is seen as 1.78 times that of the conventional ones [13]. Thus, when infectivity of virus that are currently prevalent is assumed to be unity, taking the ratio of the two, 1.35 becomes a factor. Some people say that this ratio is about 1.2, and many discussions have not yet been finalized. So, we assume to be 1.35 is the factor of L452R to N501Y virus. Table 1 shows calculation conditions. The initial value was determined so as to satisfy the above conditions. See appendix A

**Table 1.** calculation conditions

|  | **N501Y** | **L452R** |
| --- | --- | --- |
| Initial Infected People | 4400 | 5.5 |
| Infectivity | 0.09 | 0.121 |
| Sensitive Population | 500000 |  |
| Start Point | 2021/3/1 |  |
| Vaccination Start | 2021/5/15 |  |
| Vaccination Rate | 0.42%/day |  |
| Start of SOE | 2021/4/25 |  |
| End of SOE | 2021/6/20 |  |
| Time Delay until Quarantine | 14 days |  |
| Quarantine Period | 14 days |  |

SOE: State Of Emergency

As shown in the table, the effect of the vaccine is incorporated.

### 4. 2 Analysis result

#### (1) In the case of no-changing of the current measures (Case1)

Figure 1 shows the daily changes in the number of infected people. This figure indicates the pattern of the 4th wave in Tokyo. The 0th day is as of 2021/3/1. The epidemic peak is situated at the beginning of May or the end of April (fifty days or sixty days from the day of calculation start), about 800 people infected persons have been calculated. This number is roughly in line with the actual 7-day average for the 4th wave. Figure 2 shows the number of quarantined persons, including home medical treatment and hotel medical treatment. It is said that 80% of the infected person is mild person are not in hospital, then remaining of 20% are in the hospital. Therefore, at the peak more 11000 people has been quarantined and about 2200 people is considered in the hospital. According to the data of Tokyo [14] at the time of the fourth wave peak about 2400 patients are counted, then the results are consistent with the actual data. Figure 3 displays number of infected people during infection until isolation in a community. The higher this number, the higher the probability of having the next infected person. Figure 4 shows the average infection coefficient of the mutant virus, and as the infection progresses, the average value becomes closer to the infection coefficient of L452R. It shows that the L452R is becoming dominant.

**Fig. 1.**
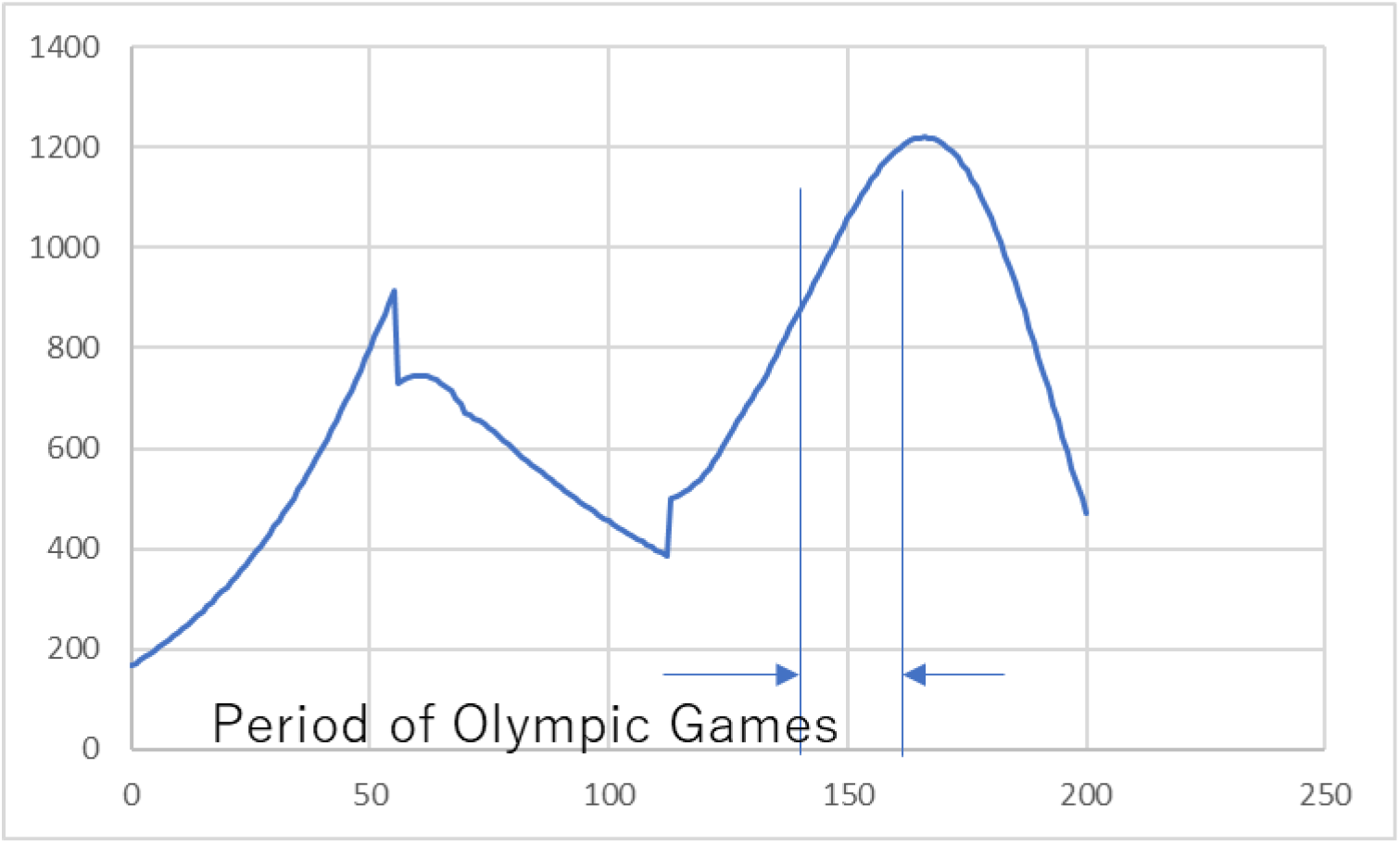
Number of daily infected people

**Fig. 2.**
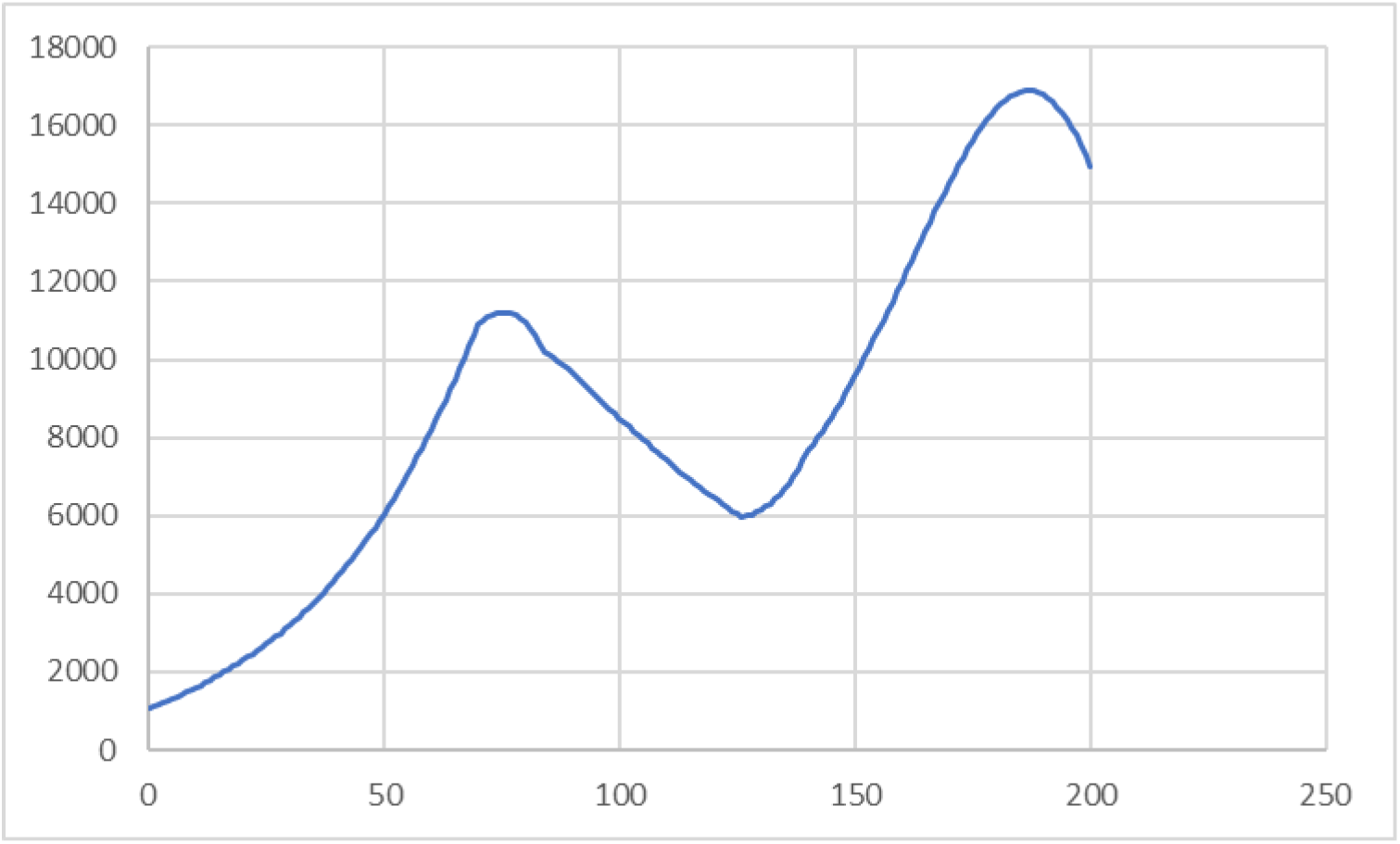
Number of quarantined Persons

**Fig. 3.**
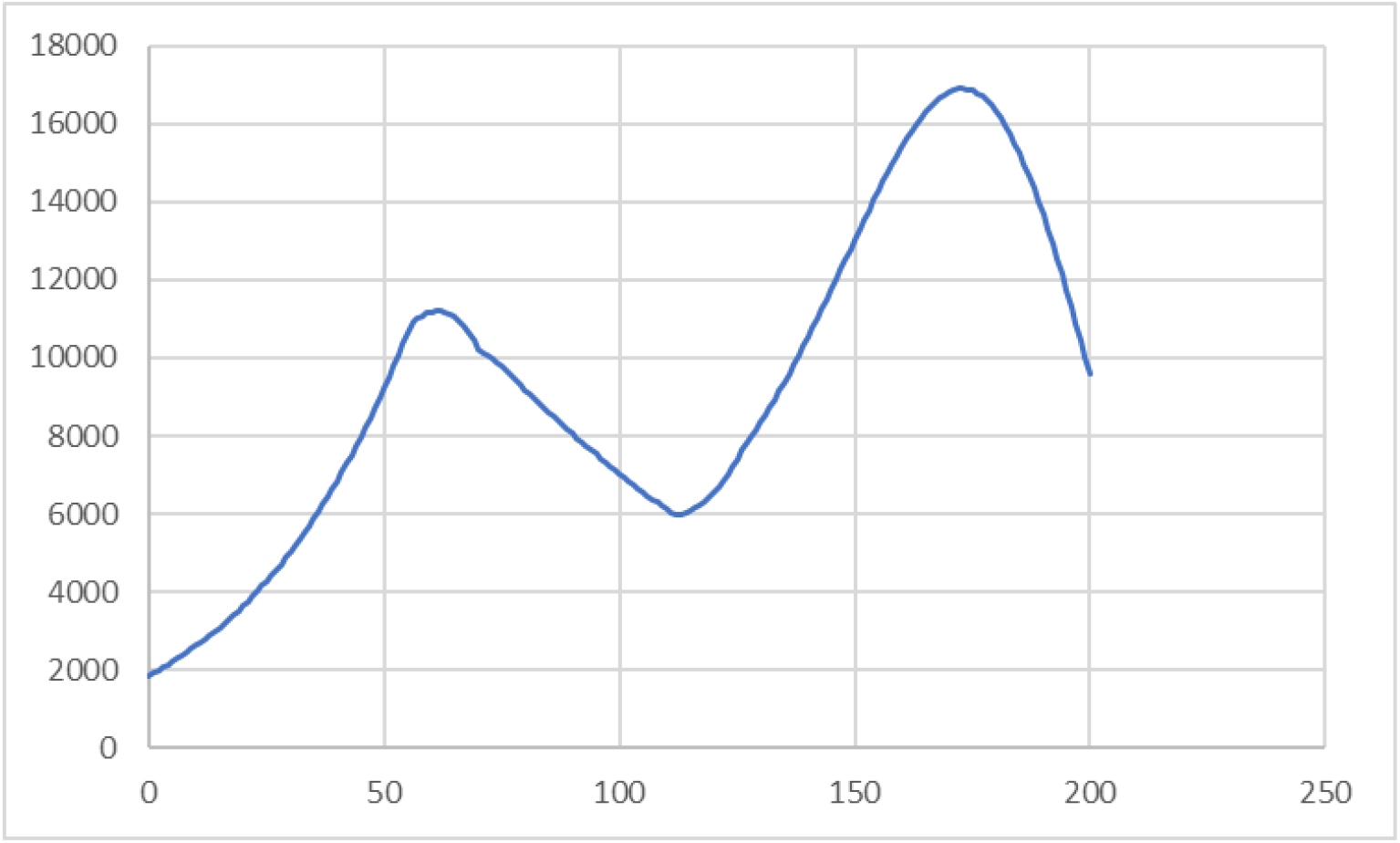
Number of infected people in community

**Fig. 4.**
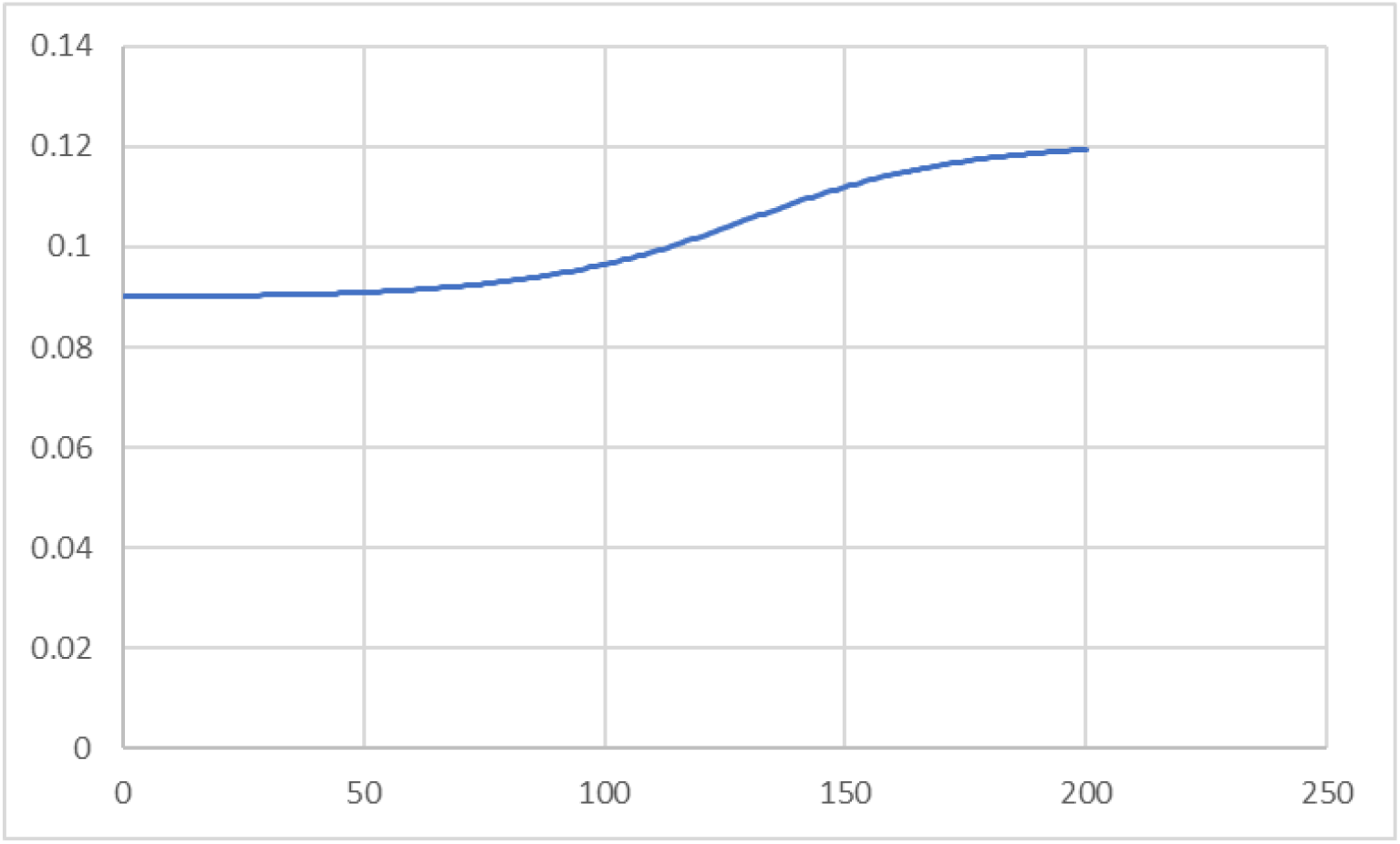
Mean infectivity

Figures 5, 6 and 7 show the number of daily infected persons, quarantined persons, and infected persons in community for each mutant virus. Blue is the number of people infected by N501Y virus, and orange is due to L452R virus. Each figure shows that the number of people infected with L452R virus has increased sharply after the end of the state of emergency. Figure 8 shows a ratio of infected persons by L452R and N501Y. The number of infected people by L452R increases from the emergency situation declared the end and becomes dominant after the point of 2021/7/6 (128 infected people number in days). In addition, from these figures, it can be seen that the infection by the highly infectious L452R rises sharply at the stage when the infection of N501Y has converged and bottomed out. As described above, this analysis also showed that the one with stronger infectivity became dominant when the infection spread.

**Fig. 5.**
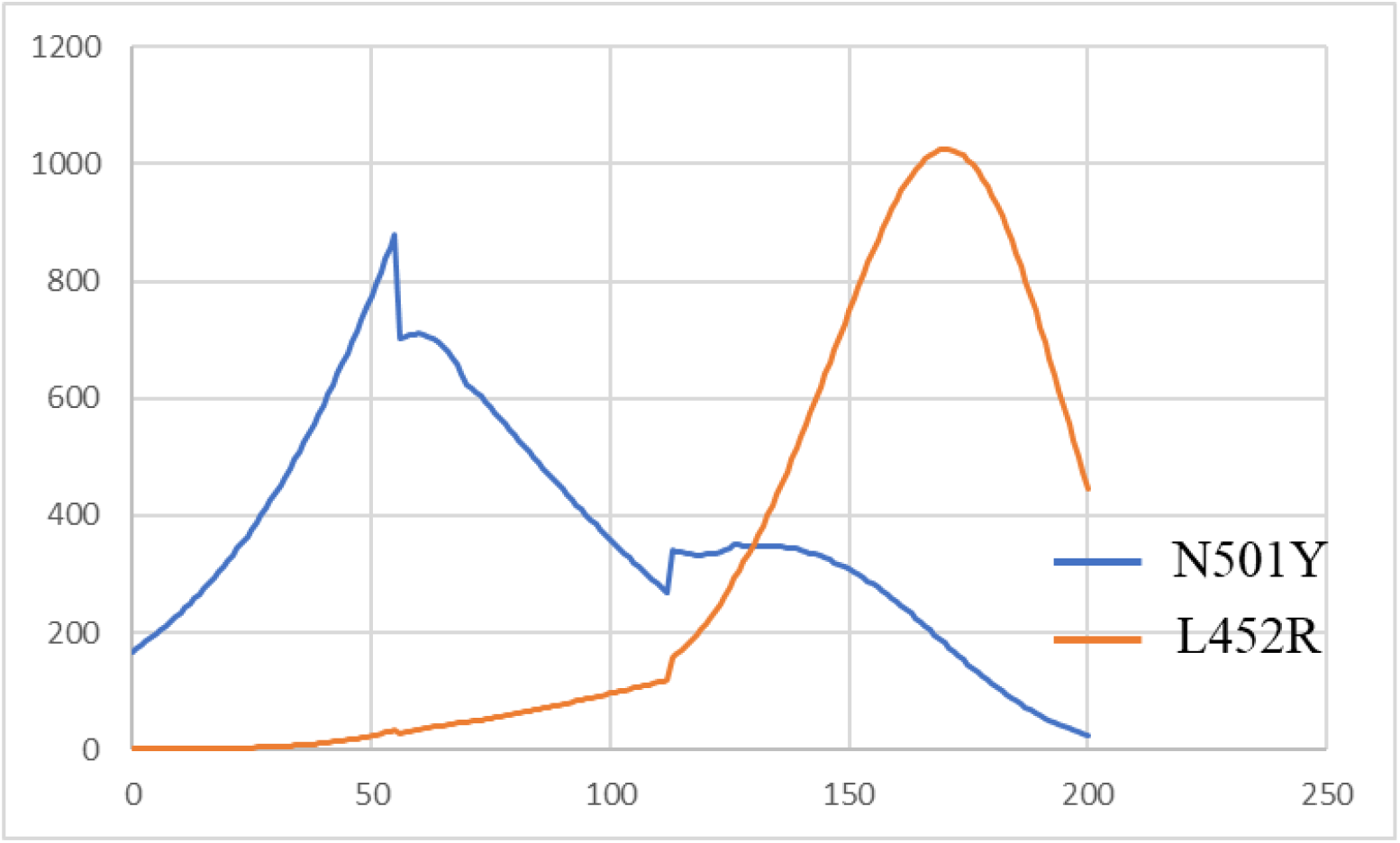
Number of daily infected people

**Fig. 6.**
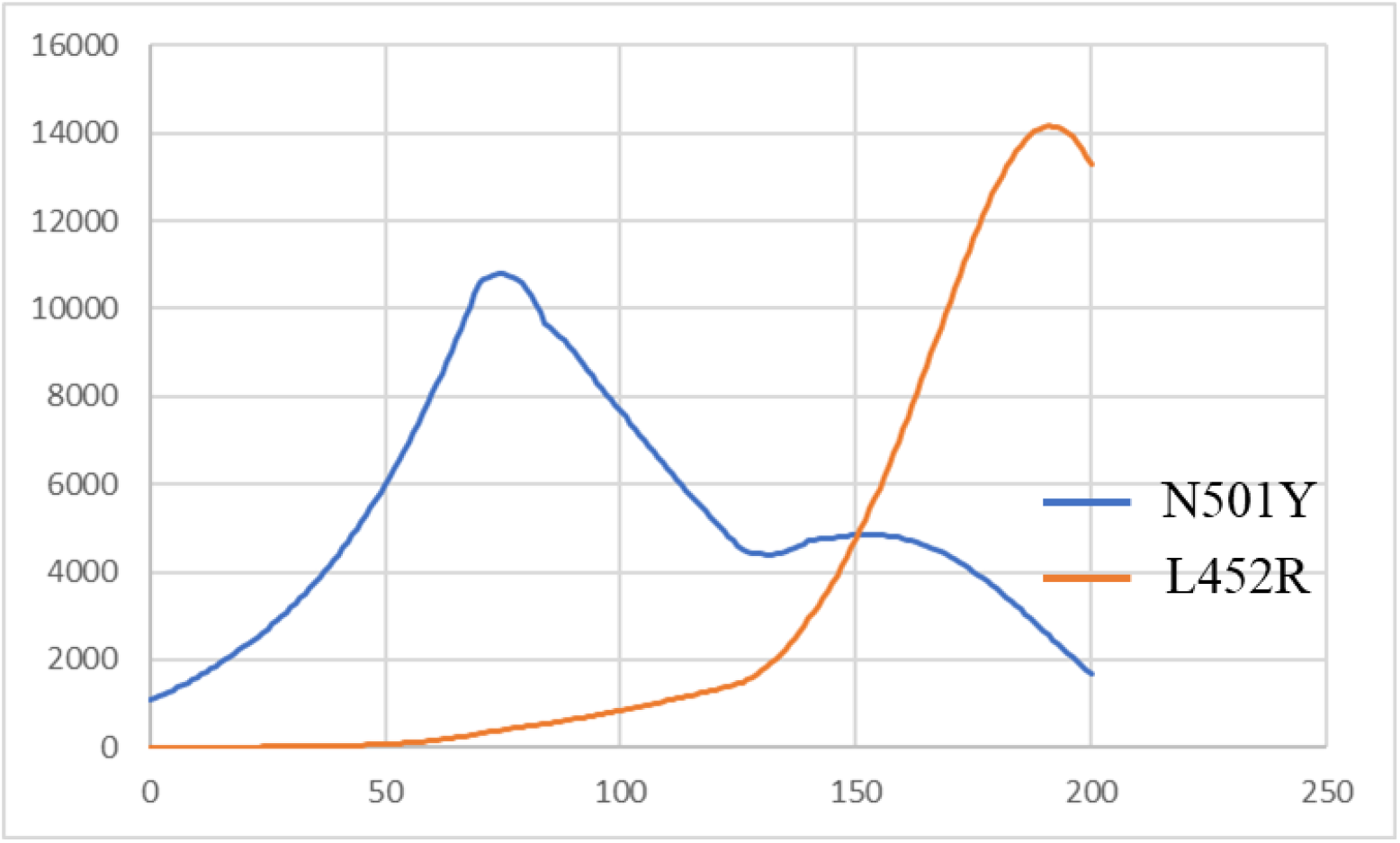
Number of quarantined persons

**Fig. 7.**
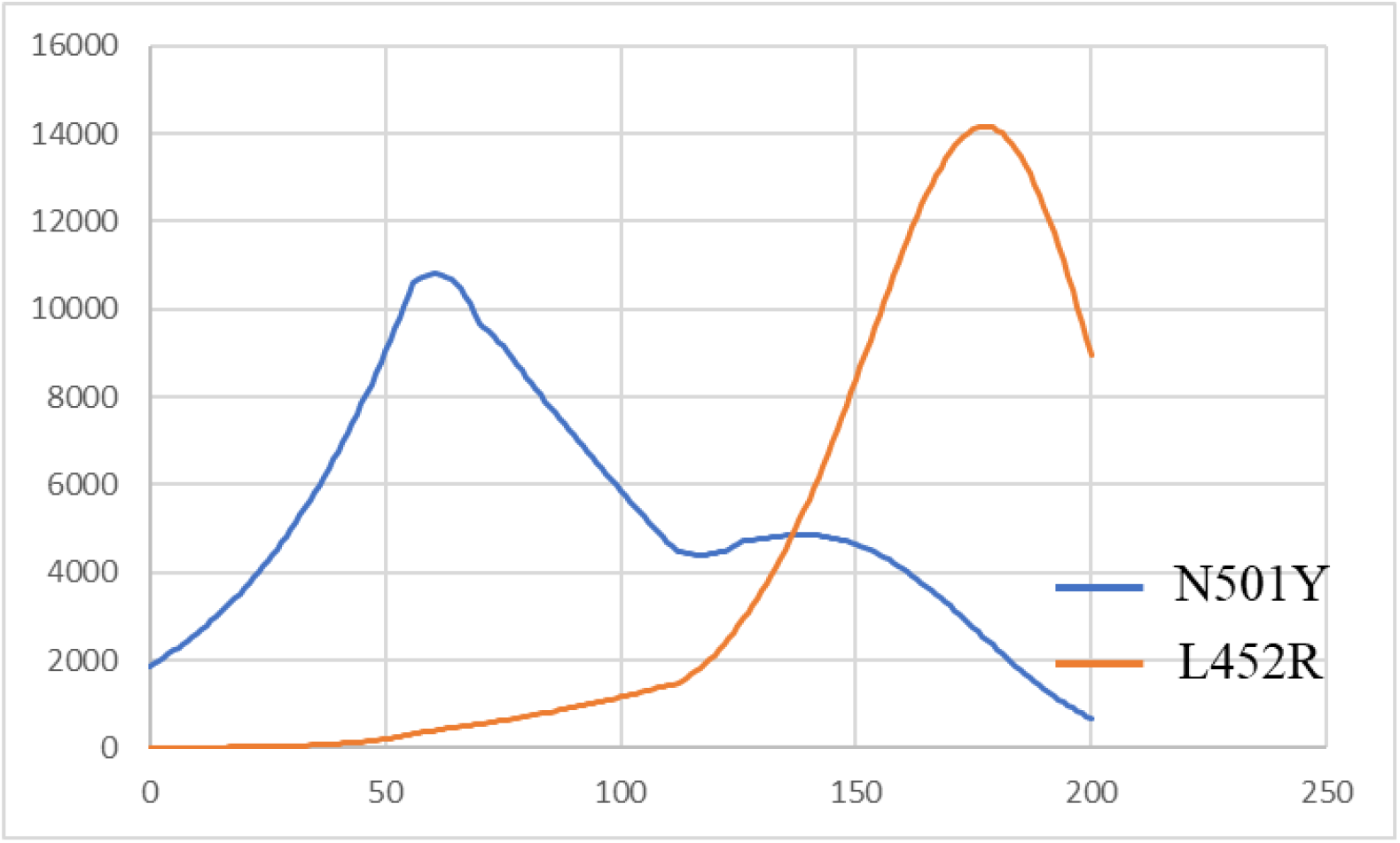
Number of infected people in community

**Fig. 8.**
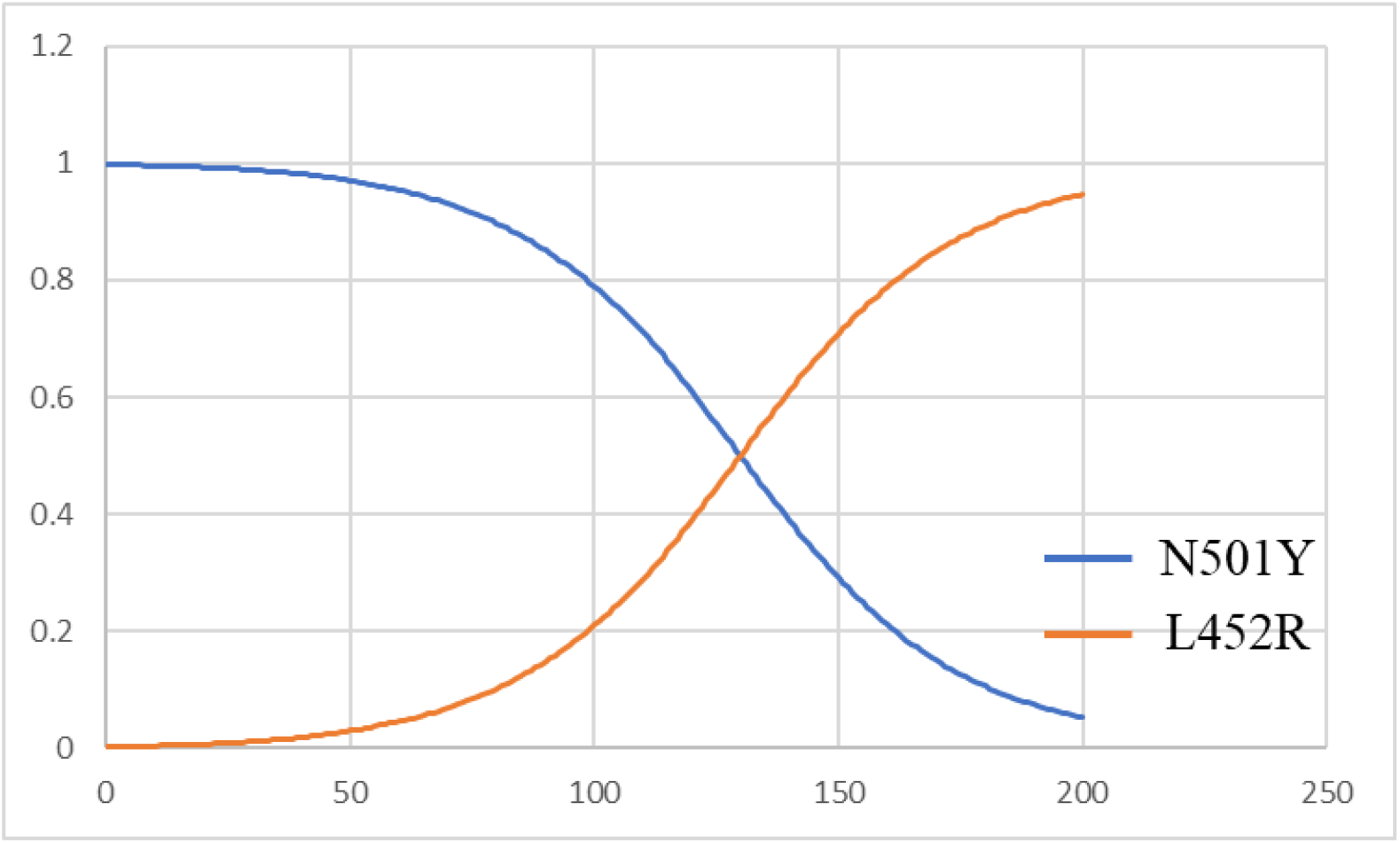
Ratio infected by N501Y and L452R

#### (2) Measure 1: In the case of extending the state of emergency to 6/30 (Case 2)

Figure 9 shows the transition of infected persons when the state of emergency is extended to 6/30. Peak of infected persons 100 decreases as people, but the big improvement of the infection situation is not observed. Therefore, the extension until 6/30 has no effect on suppressing infection.

**Fig. 9.**
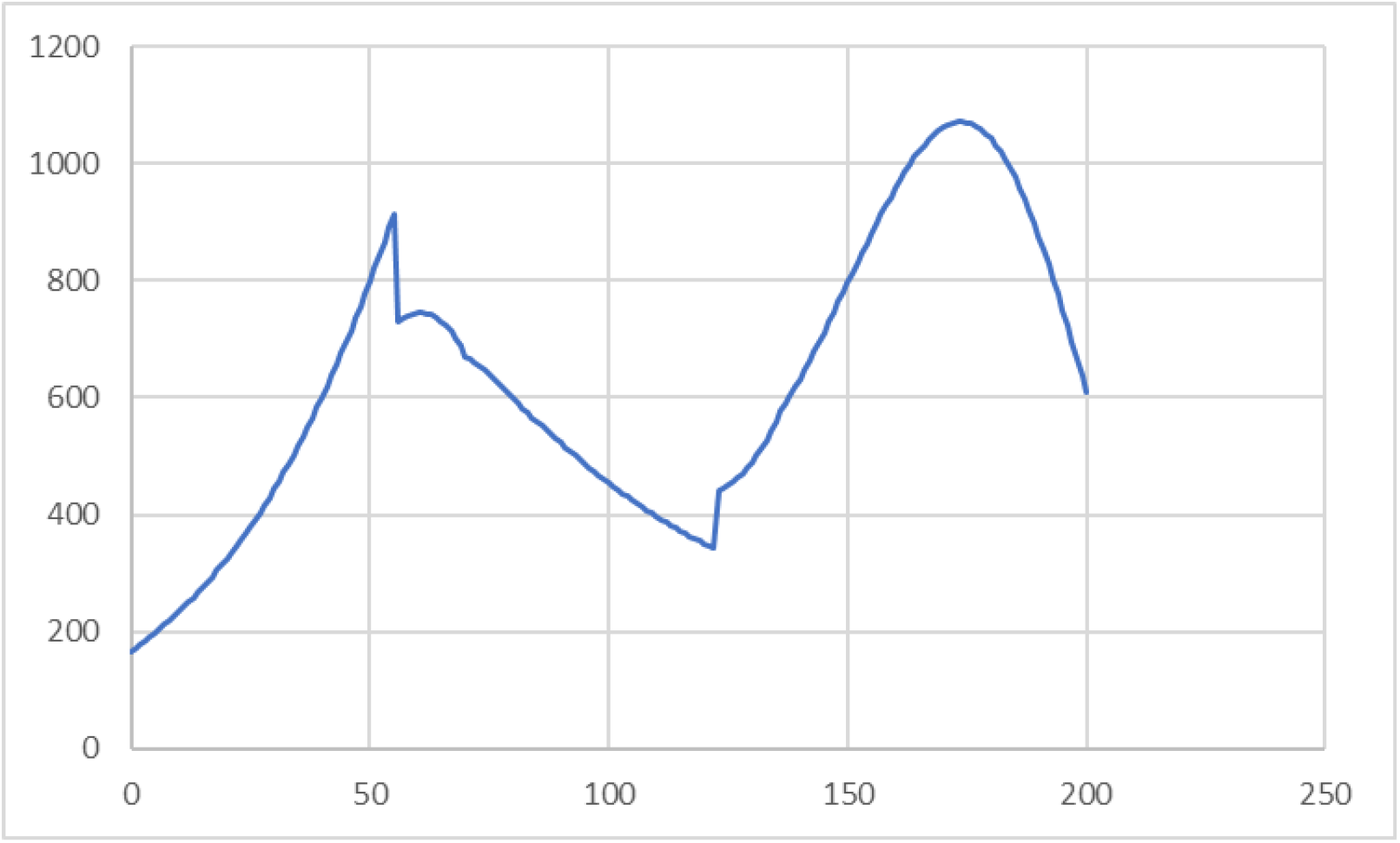
Number of daily infected people in Case 2

#### (3) Measure 2: To shorten the period from infection to isolation by PCR test or antigen test (Case 3)

This measure is not effective unless as many people as possible participate. It is good if you know that you will be infected, but usually you do not know, so you need to have many people check it regularly. For that purpose, many people can participate by issuing a negative certificate for a limited time (up to one week) in the test and confirming it at restaurants and event venues. In this way, it is possible to shorten the period from infection to isolation for two days or one day. In this study, this period is set to 14 days due to the consistency of the data, but it is estimated to be about 7 days in reality. Then, one actual day short is equivalent to two days of the study. We set the period to 12 days. Considering five days as preparation period after the end of emergency, the implementation date was set to 6/25. The results are shown in Fig. 10. As shown in the figure, the spread of infection after the peak of the 4th wave is suppressed to about 600 people. Searching for infected people early and shortening the infection period in this way is the most effective method other than vaccines as an infection control measure.

**Fig. 10.**
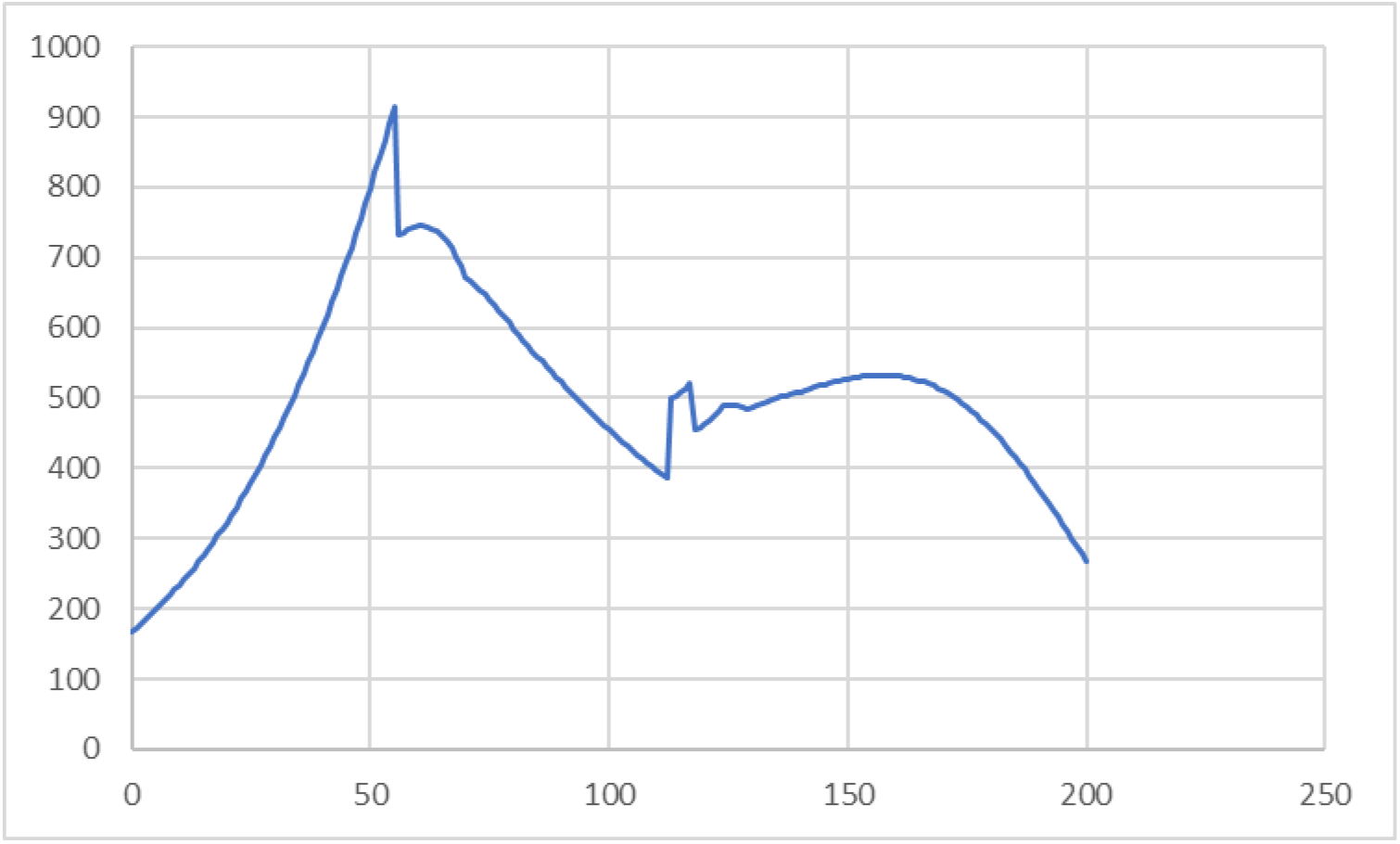
Number of daily infected people in Case 3

In Fig. 11, the infection status of each cases are plotted together. The horizontal axis is the date from 2021/3/1. The blue line shows the infection status in Tokyo on a 7-day average [14]. From this figure, it can be seen that Case 3 suppresses the spread of infection most and is effective.

**Fig. 11.**
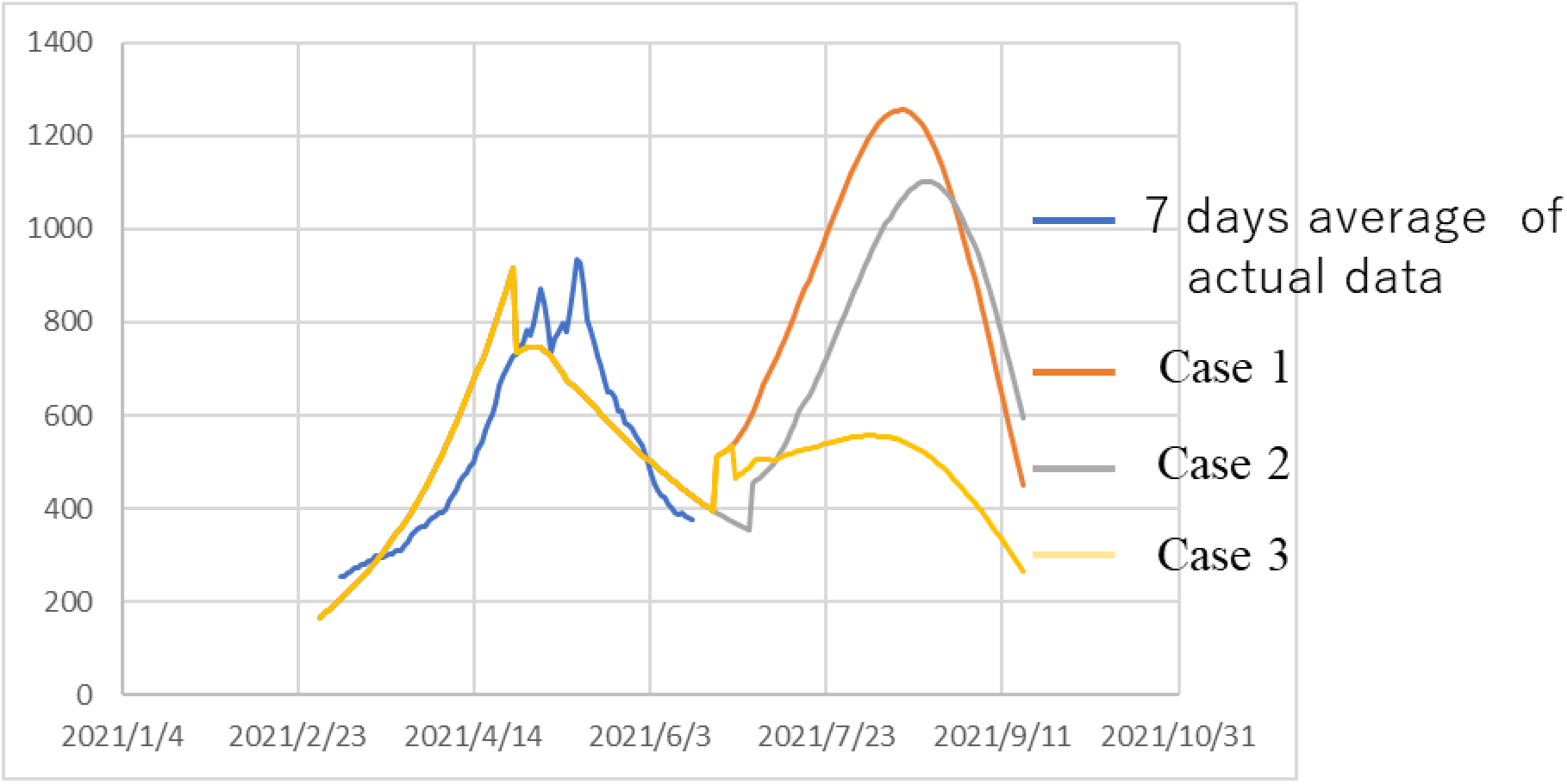
infection status in each case

## 5. CONCLUDING REMARKS

We have extended the ATLM which has been developed to simulate the status of infection with various mutant viruses. The developed model was applied to the 4th wave of Tokyo and the following results were obtained.

1. The fourth wave will bottom out near the end of the state of emergency, and the number of infected people will increase again.
2. The rate of increase will be mainly by L452R virus, while the increase in N501Y virus will be suppressed.
3. It is anticipated that the infection will spread during the Olympic Games.
4. When mutant virus competes, the infection of highly infectious rises sharply while the infection by weak infectivity ones has converged.
5. It is effective as an infection control measure to find an infected person early and shorten the period from infection to quarantine by PCR test or antigen test as a measure other than vaccine.

## Data Availability

We used time-series data of COVID-19 for March 1 through June 10, 2021 in Tokyo, which is publicly available.

https://stopcovid19.metro.tokyo.lg.jp/en

## DATA AVAILABILITY

We used time-series data of COVID-19 for March 1 through June 10, 2021 in Tokyo [14].

## AUTHOR CONTRIBUTIONS

M. Koizumi developed the epidemiological model. M. Utamura verified the numerical results. S. Kirikami identified parameter values from data. All authors have read and agreed to the published version of manuscript.

## ACKNOWLEDGMENTS

MK is the former researcher of Hitachi Ltd., MU is the former professor of Tokyo Institute of Technology and SK is the former engineer of Hitachi Ltd.

## APPENDIX A

Calculation of the first approximation of M and α

In ATLM, the rate of infections (daily new cases) 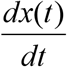 is given by the following equation with the collective population M and the transmission rate α.

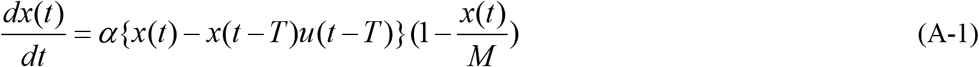

Here, *x*(*t*) : cumulative infections, *T*: delay time from infection to quarantine, u: the step function.

Since daily new cases varies significantly, integration is used to smooth them. The intervals[*t*_1_, *t*_2_],[*t*_3_, *t*_4_](0 < *t*_1_ < *t*_2_ < *t*_3_ < *t*_4_) are used as those of interest.

The following equation is obtained from the integration of (A-1) in the interval[*t*_1_, *t*_2_].

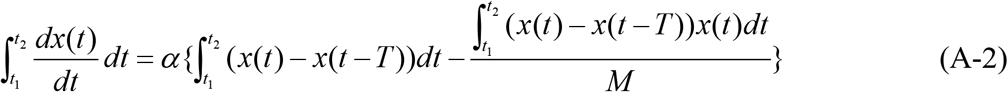

Normally, *T* < *t*_1_. For this reason, we set the step function u = 1.

(A-2) is modified to obtain (A-3).

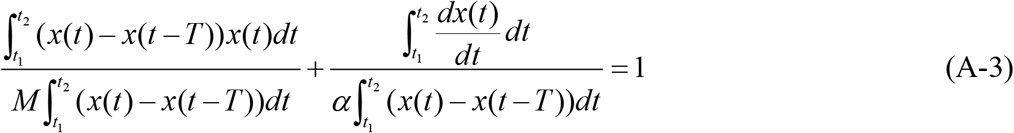

Similarly, (A-4) is obtained in the interval[*t*_3_, *t*_4_].

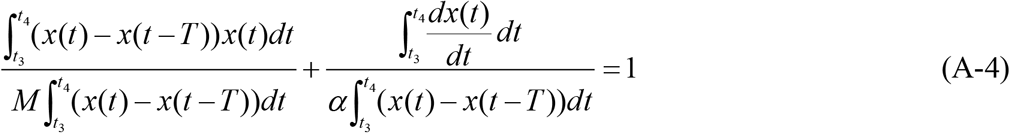

(A-3) and (A-4) are linear equations of 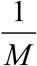 and 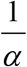. Then M and α are calculated.

Usually, the following inequality holds in the range of monotonously increasing 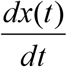.

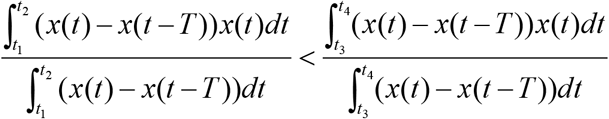

The condition (A-5) is sufficient for M and α to be positive.

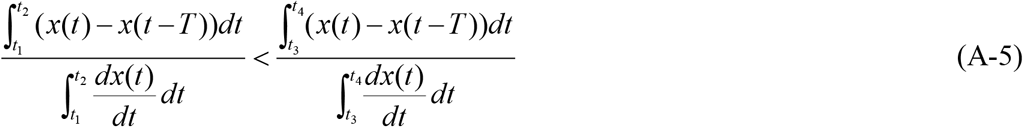

The case of Tokyo

The source is from the following. Updates on COVID-19 in Tokyo | Tokyo Metropolitan Government COVID-19Information Website

The intervals were 2021 3/22 - 4/4 (14days) and 4/5 - 4/18 (14days) before the state of emergency. The delay time from infection to quarantine was T = 14 days. Therefore, the measured number of daily new cases 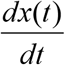 on the day *t* was used as the number of daily new ones T = 14 days ago.

The cumulative infections *x*(*t*) were accumulated from the minimum number of daily new cases on February 15th.

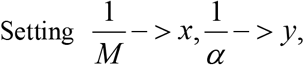

we obtained the following two linear equations.

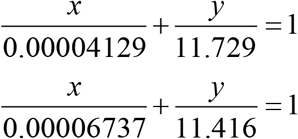

From these equations, M = 3.66013 * 10 ^ 5 and α = 0.0913 were obtained as the first approximations.

Integration was calculated with the trapezoidal formula. It was also used for the number of daily new cases 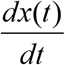. Therefore, numerically,

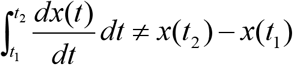

In the above, the calculation of the first approximations of M and α is shown by taking the case of Tokyo. These must be so tuned that they are consistent with the other measurements.

